# Guiding caregivers through disclosure: a qualitative investigation into caregivers’ responses to a disclosure intervention for HIV-infected children in Soweto, South Africa

**DOI:** 10.1101/2021.10.14.21264620

**Authors:** Celeste Joyce, Candice Ramsammy, Lisa Galvin, Given Leshabane, Afaaf Liberty, Kennedy Otwombe, Janice Buckley, Minja Milovanovic, Avy Violari

**Affiliations:** Perinantal Human Immunodeficiency Virus Research Unit, Faculty of Health Sciences, University of the Witwatersrand, Johannesburg, South Africa; School of Public Health, Faculty of Health Sciences, University of the Witwatersrand, Johannesburg, South Africa; African Potential Management Consultancy, Kyalami, South Africa; Department of Psychiatry, University of the Witwatersrand, Johannesburg, South Africa

**Keywords:** HIV, disclosure, stigma, children, caregivers

## Abstract

Awareness of Human Immunodeficiency Virus (HIV) status improves health outcomes in HIV infected children, yet disclosure is often delayed by hesitant caregivers. This qualitative investigation explored how 30 caregivers responded to a HIV Disclosure study conducted between 2017 and 2020 at Chris Hani Baragwanath Academic Hospital, Soweto, South Africa. Caregivers were assisted in disclosing to their children, aged 7-13 years; followed by a sub-sample of caregivers interviewed to elaborate on findings.1) Barriers to disclosure included caregivers being ill-equipped, fear of negative consequences and children considered lacking emotional or cognitive readiness. 2) Deflecting diagnosis and the need for medication motivated caregivers to disclosure. 3) Apprehension was evident during disclosure, however, overall disclosure was a positive experience with the support of the healthcare providers. These results highlight the significant role healthcare providers play in supporting caregivers through the disclosure process and how, in turn, this has a positive impact on the community as a whole.

## Introduction

The increase in Antiretroviral Therapy (ART) coverage led to a decline in HIV related deaths of children in Southern Africa, now making Human Immunodeficiency Virus (HIV) a chronic condition that requires comprehensive multidisciplinary care (1-10). Disclosing HIV status to perinatally-infected children remains a challenge despite evidence suggesting that it improves health and emotional outcomes (2, 3, 8-15). The SADoH (16) suggests that partial disclosure takes place from 3 years to 9 years of age, and full disclosure is to take place from 10 years of age. Furthermore, this process should be followed-up. The WHO (4) suggests disclosing to school going children between 6 and 12 years of age (16). The World Health Organisation (WHO) and the South African National Department of Health (SANDoH) encourage a comprehensive approach to disclosure which ensures the child’s physical, emotional, cognitive and social well-being is safeguarded by pre-disclosure not being completed in front of the child and children of school going age to be made aware of their caregivers’ HIV status (4, 10, 11, 13, 16). Despite this, studies have shown that many children are unaware of their HIV status and that their caregivers are hesitant to disclose to them (17).

Disclosure has far reaching psychosocial implications (18). Caregivers and their HIV-infected children live as a part of a greater “whole”. In accordance with Social Learning Theory, an individual is a member of his/her immediate family, extended family, culture, peer group, school and country. Not only do they influence their environment but they are directly influenced by the attitudes, perspectives and behaviours of others (19, 20). Caregiver attitudes towards HIV and disclosure are shaped by their personal experiences within the environment and the attitudes of those around them. This, in turn, shapes when and how disclosure is addressed in their own families (15, 20, 21). The literature has identified several barriers to disclosure including: social stigma; fear of discrimination; the impact on the child’s emotional/psychological well-being; fear of the child’s resentment; untimely disclosure of HIV status to others; exposure of the caregiver’s “personal secrets”; and caregiver’s lack of knowledge on how and when to approach the disclosure questions (4, 13, 16, 17, 22-27). A recent study in Ghana (28) reported that caregivers lacked knowledge on how to disclose to their children, and that this poor knowledge was the main reason for delayed disclosure.

Few mothers were reported as being confident in their ability to disclose their status due to social stigma and the possibility of this stigma being passed onto their children (21). Mothers more effective in disclosing their own statuses had better relationships with their children and this in turn improved the children’s adjustment to the new knowledge of their mother’s HIV status (29, 30). The VUKA study, which focused on pre-adolescent children awareness of their HIV status and family members, emphasised the importance of family involvement in mental health and health promotion interventions (31). Improvements in mental health, behaviour, HIV treatment knowledge and medication adherence, stigma and communication between the child and family members involved were noted (31).Therefore, interventions aimed at assisting mothers disclose their child’s HIV status to their children in a developmentally appropriate manner may also improve the mother’s confidence in disclosing her own status.

We developed a study that guided caregivers through disclosure and explored their responses and interactions with their children before and after disclosure, and psychometric investigations of the impact of the disclosure process on the child’s emotional state and adaptive behaviour (29). This 78-week study used disclosure material (30-32) to standardise the process across all participants, with both the caregivers and healthcare providers directly involved.

Here we present the caregivers’ motivation for and response to each stage of the disclosure process. As the caregiver is at the core of the disclosure or non-disclosure issue, it is important to understand their responses and perceptions and use these to shape future interventions.

## Methods

### Study Setting and Participants

The disclosure study was conducted between 2017 and 2020 at the Perinatal HIV Research Unit (PHRU), Chris Hani Baragwanath Academic Hospital, Soweto, South Africa. In keeping with both the SANDoH and WHO guidelines, children between the ages 7-13 years, who had not been previously fully disclosed to were recruited along with their caregivers. Caregivers who attended PHRU clinics with their children were approached and children, who were not disclosed to or had received partial disclosure, and their caregivers were invited to participate in the study. During the first study session the caregivers were seen without the child participants and provided with information about the study process. The participating caregivers provided consent and following this the child participants were asked to provide assent (32). Thirty caregivers (i.e. the child’s biological parent, legal guardian, foster parent, or another person that was responsible for the protection and promotion of the child’s health, well-being and development) participated in the main study. Following the completion of the main study in 2020, a sub-sample of 15 caregivers were invited to participate in the in-depth interviews where they shared their perspective of the disclosure program. These participants were purposely sampled to ensure the capture of a broad range of experiences.

### Study Procedures

Study visits took place over 78 weeks, alternating between disclosure counselling sessions (completed week 72) and psychometric sessions (completed week 78), six weekly. The counselling sessions were led by healthcare providers, trained by the study Principle Investigator to use the “Right to Care Mini Flipster” (33) and the Joint United Nations Programme on HIV and AIDS (UNAIDS) (34) disclosure material. The initial pre-disclosure session was attended by the caregiver and the healthcare provider. During this session the caregiver was provided with more information and prepared for the disclosure process. The child and the caregiver participated together in the sessions that followed, facilitated by the healthcare provider. The healthcare provider made notes about the process during each session.

The initial sessions provided education about health, the immune system and the importance of medication adherence. The following sessions then assessed the child’s knowledge about HIV and unclear concepts were revised. Based on the child’s comprehension of these sessions, full disclosure was done by the caregiver with the support of the healthcare provider. Disclosure counselling sessions focused on children’s feelings, made them aware of their support structures, and emphasised their own agency in their care. The process was individualised and flexible, allowing for additional visits, revisiting of topics, and accelerating or decelerating the process.

Following the completion of the main study (between nine and 15 months after full-disclosure), a subset of 15 caregivers were invited to participate in the in-depth, structured interviews aiming to better understand their experiences of the disclosure study. Purposeful sampling was used to ensure a wider age range, both female and male perspectives, as well as the perspectives of one caregiver who discontinued the study. Two participants discontinued the study, both never receiving full disclosure. One child’s caregiver was no longer contactable after week 24. The other child’s caregiver was unavailable after week 48 but did consent to participating in the in-depth interviews.

Consent was obtained and in-depth interviews were conducted with 15 caregivers. Interviewers were proficient in counselling and conducting interviews. They received training on the interview schedule and were fluent in the spoken local languages of the participants and English. As they were also familiar with the participants’ culture, it was unlikely that cultural barriers would have skewed participant answers.

### Tools

#### Counselling Tools

This study provided developmentally appropriate disclosure counselling using the “Right to Care” Disclosure Tool and the United States Agency for International Development (USAID) / U.S. President’s Emergency Plan for AIDS Relief (PEPFAR) Disclosure booklets were used to standardise the disclosure process. The “Right to Care” Disclosure tool (33) consists of two developmentally appropriate flipster books: “Preparing Children for Pre-Disclosure Through Play” utilises stories, activities and songs to teach the younger child, below 12 years of age, about germs, healthy living and illness; and “Mini-flipster: Disclosure Tool for Adolescents Ages 12 & Up” explores the topics of Pre-Disclosure, Full-Disclosure and Post-Disclosure using the Socratic method of questioning. These tools were used together and developed to assist caregivers in the disclosure process.

#### Disclosure Counselling Form (DCF)

The DCFs were specifically developed by the research team for this study to record disclosure progression, provide progress notes and information regarding who accompanied the child. After each counselling session, the healthcare provider completed a DCF; recording their observations regarding the caregiver’s and the dyads’ interaction, and information provided by the caregiver during the session. The DCF was used to maintain structure and was also introduced to identify any psycho-social difficulties which could be addressed immediately.

### Data Collection

#### Healthcare provider’s observations of the counselling sessions

The healthcare providers (a social worker, counsellor or nurse) noted their observations on the DCF after each disclosure counselling session for each participant (approximately 6 visits). This qualitative component relied on the healthcare providers’ detailed observations of the caregivers’ and the children’s responses and behaviour during the counselling sessions. Any other relevant information that arose on the day was also recorded on this form.

The DCFs provided healthcare provider perspectives and observations throughout each stage of the disclosure process. There is little information in the literature regarding caregivers’ reactions to the disclosure process and it was felt to be a valuable addition to the area of disclosure. As this could be considered to be subjective and from the perspective of the healthcare provider it was felt that it would be pertinent to include the in-depth interviews in this study as they explored specific themes more deeply providing a richness to this study.

#### In-Depth Interviews

An in-depth, structured interview guide was developed by the research team. Open-ended questions were used to elicit the perspectives of the caregivers on the disclosure process. The interviews were developed and formulated with the purpose of understanding the challenges and concerns caregivers had around disclosing to their children before joining the disclosure study; the disclosure process itself; and to identify any concerns and parenting challenges that may arise in the future. These interviews were conducted by a registered counsellor and research psychologist. The registered counsellor did the psychometric assessments with the participants and had minimal contact with the caregivers. The research psychologist was not involved in the main study. The interviews took approximately 30-40 minutes to complete. Caregivers were interviewed primarily in English. However, questions were clarified and participants were able to respond in their language of preference (English, isiZulu or seSotho) and with their consent, audio recordings were made. The recordings were later transcribed and all transcriptions were translated into English for analysis.

### Data Analysis

Information regarding barriers to disclosure, as well as observations relating to the caregiver and their response to the intervention from both the DCF’s and in-depth interview transcriptions were analysed manually using content analysis. The analytic approach included the development of a code book, reviewing of the codes, coding and interpreting the data in order to describe, compare, categorise and conceptualise (35, 36).

Four analysists (a Clinical Psychologist, Psychiatrist, Research Psychologist and Registered Counsellor) independently reviewed and familiarised themselves with copies of the DCFs and interview transcripts. The analysts represented different cultural backgrounds and ages, and counselling fields and areas of expertise. Thus being experienced in interpreting subtle emotional nuances in responses and ensuring trustworthiness, credibility and reliability in the data interpretation. This assisted with improving the likelihood that cultural or training bias would unduly influence perceptions of results when coding data. Furthermore, none of the analysts were counsellors in the study, reducing the likelihood of bias when interpreting the data.

Initially, analysts working individually identified main ideas (meaning units) within the DCFs. These identified meaning units were discussed in the group and once agreed upon, the code book was developed. The data was then coded and codes of similar content or context were grouped together. These codes and categories were again compared across analysts, merging common codes to formulate a single tabulation of categories. Discrepancies were discussed until consensus was reached. The categories were used to re-evaluate the data and to quantify the frequency of categories noted. Themes were then identified and the categories were grouped within the themes. The in-depth interview transcripts were treated independently, developing a separate code book and coding the in-depth interviews after the analysis of the DCFs had been completed. Similarities and differences between the two sets of data were identified and combined to provide a more complete understanding of the caregivers’ reasons for not disclosing to their children before joining the program.

Caregiver demographics were analysed using descriptive statistics.

### Ethics/Protection of Human Subjects

Ethical approval to conduct this study was obtained from the University of the Witwatersrand Human Research Ethics Committee (HREC, reference number 170107), Johannesburg. Informed consent was obtained from all caregivers and assent was obtained from all child participants in the Disclosure Study.

## Results

### Study Population (See Table 1)

Thirty HIV positive child and caregiver dyads were enrolled into the study The median age of the caregivers were 39.05 (IQR = 34-49). The majority of the caregivers (90%, 27/30) were biological parents and predominantly mothers (24 mothers vs 3 fathers), and the remaining 3 were grandmothers or aunts. However, during the study one father was unable to continue and the child was then accompanied by their mother; a second participant’s mother was unable to complete the study and the father attended the final counselling session. In both cases this was due to employment commitments. Twenty-eight caregivers were HIV-infected themselves and 27 were receiving ART. The majority of the adults were unemployed (67%) and 83% a secondary level of education.

**Table 1:** Caregiver Demographics.

|  |  |  |
| --- | --- | --- |
| **Relationship to participant** | Mother (%) | 23 (76.66) |
|  | Father (%) | 4 (13.33) |
|  | Other (%) | 3 (10.0) |
| **HIV Status** | Positive (%) | 28 (93.33) |
|  | Negative (%) | 2 (6.67) |
| **Receiving ART** | Yes (%) | 27 (96.43) |
|  | No (%) | 1 (3.57) |
| **Education Level** | Post Grade 12 (%) | 3 (10.0) |
|  | Secondary (%) | 1 (3.33) |
|  | Primary (%) | 25 (85.33) |
|  | Unknown (%) | 1 (3.33) |
| **Employment Status** | Employed (%) | 10 (33.33) |
|  | Unemployed (%) | 20 (66.67) |

Of the 15 caregivers who participated in the in-depth interviews, the majority were mothers (13 mothers vs 2 fathers) with a median age of 38,08 years (IQR = 34-43). Thirteen of 15 caregivers had fully disclosed to their children by the end of the study. One participant completed all study visits but due to their young age and insufficient concept comprehension, the participant did not receive full disclosure. Moreover, the remaining dyad discontinued the study prior to full disclosure.

Three central themes, related to the caregivers’ experience of disclosure, emerged during analysis: their barriers towards disclosure; their reason for disclosing through the study; and the caregivers’ response to disclosure.

### 1. The caregivers’ reservations to disclosure of the child’s HIV status (See Table 2)

Caregivers were asked at the first disclosure counselling session what had prevented them from disclosing their child’s HIV status to them. This open-ended question, recorded on the DCFs, was identified as a central theme during the content analysis. These reasons were further supported by the in-depth interviews. Three sub-themes were identified:

**Table 2:**
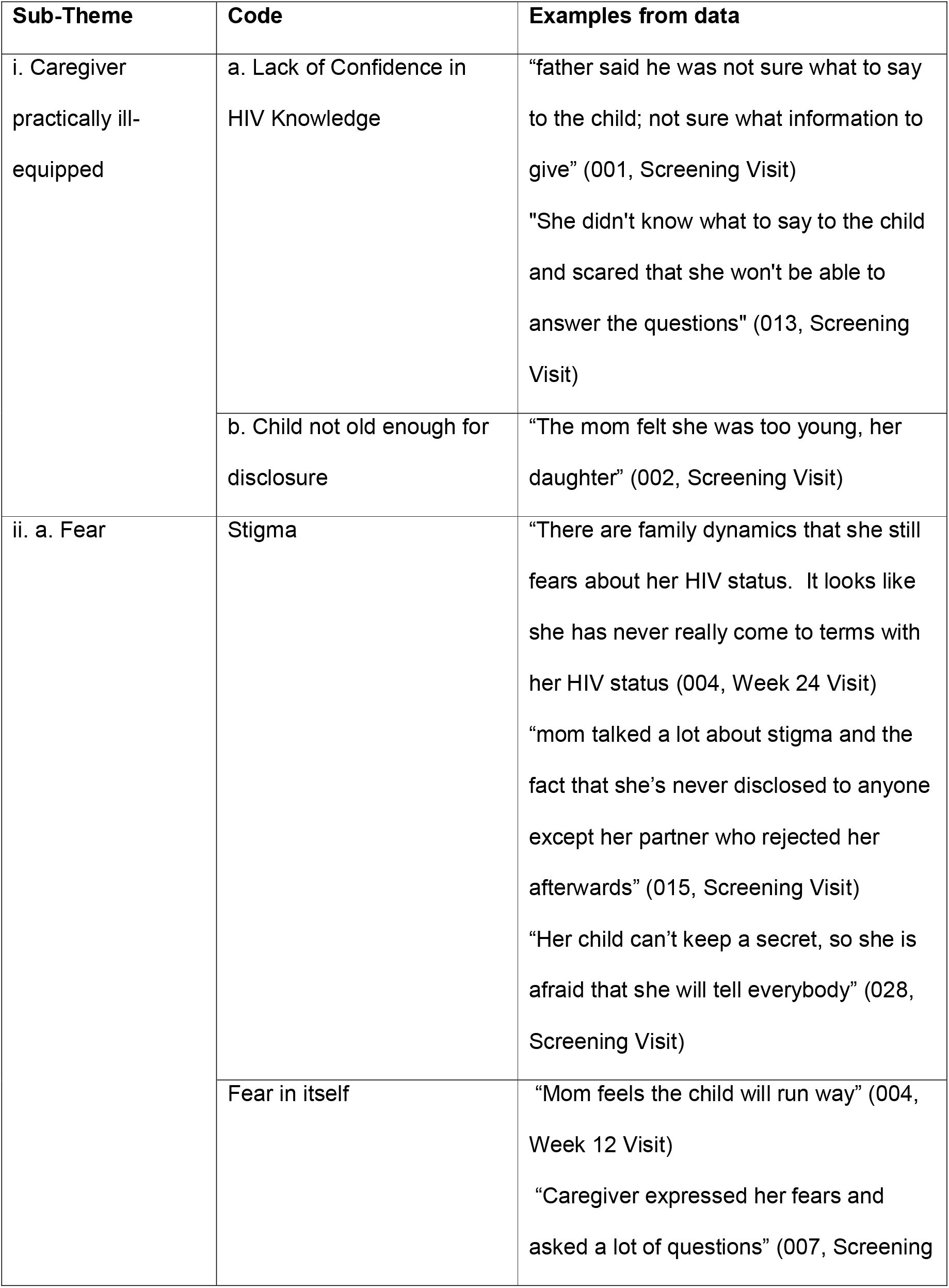
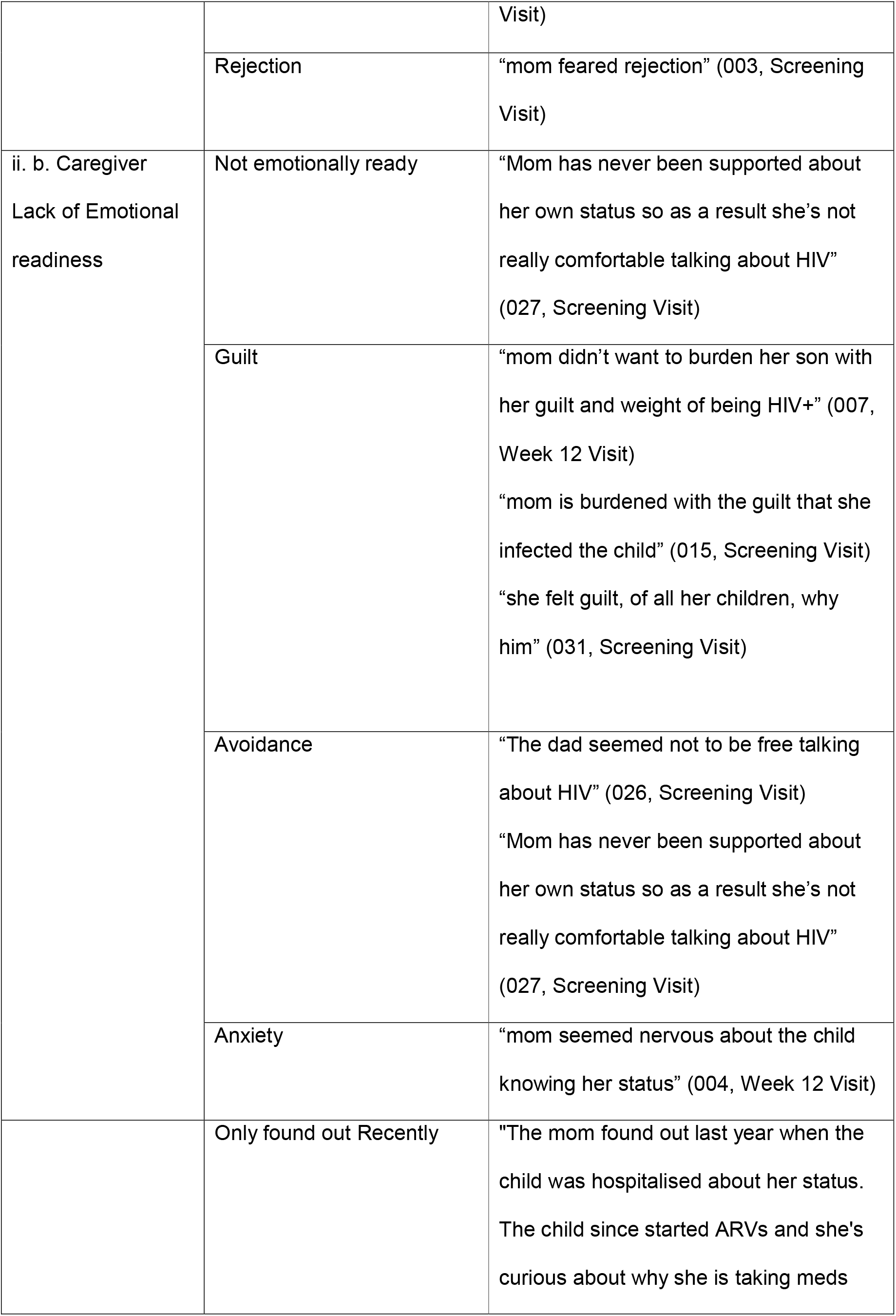
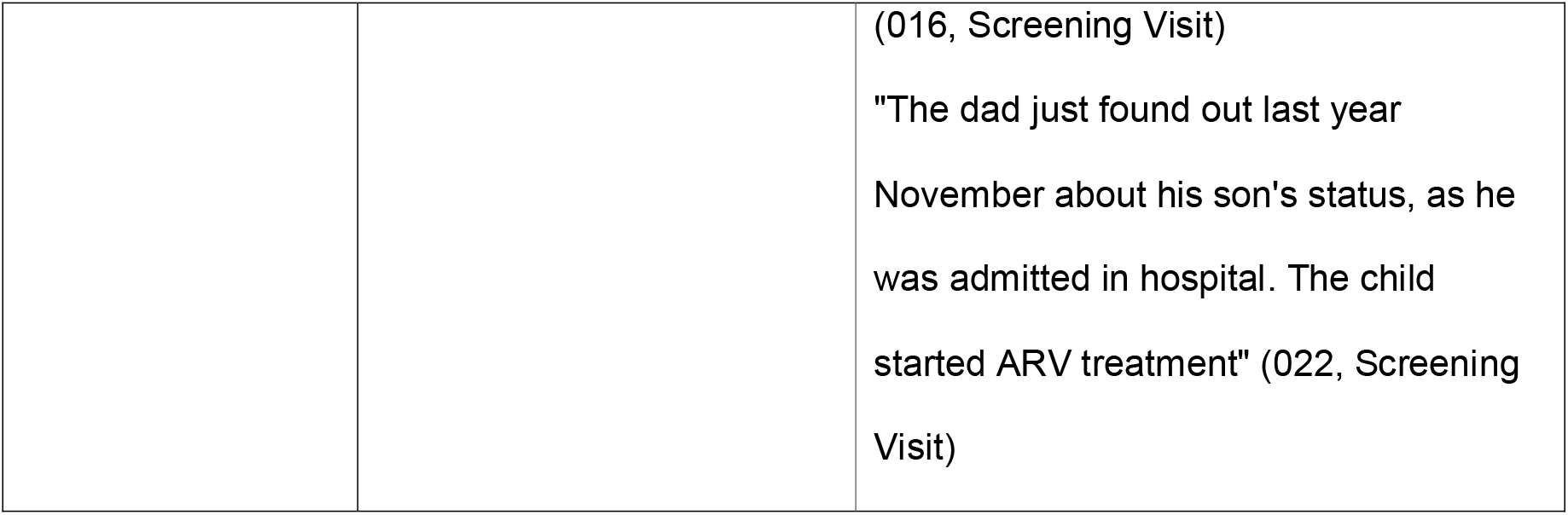
Barriers to Disclosure taken from the Disclosure Counselling Forms.

1.1) Caregiver practically ill-equipped to disclose
1.2) Fear
1.3) Lack of emotional readiness

#### 1.1 Caregiver practically ill-equipped for disclosure

The most common reason for not disclosing (43%, 13/30 caregivers) was that they felt that the children were still too young and that younger children would not be able to understand HIV/AIDS. There was, however, no reference to a particular age at which the child would be “ready” or why they thought the child wouldn’t understand the information. Although the caregivers indicated that the participants were too young to undergo disclosure, this was better associated with the caregivers’ own insecurities and inadequacies in their ability to disclose to their children. During the interviews this was explored and caregivers gave age ranges between 11-18 years as an appropriate age for disclosure as they would have a better understanding of seriousness of HIV and the implications for their lives.

Almost three quarters (70%, 24/30) of caregivers felt ill-equipped to disclose, citing challenges in communicating with their children about their illness. Eleven caregivers (37%, 11/30) reported a lack of confidence in their own HIV knowledge restricted their ability to disclose to their children. Some caregivers did not know what and how much information to share, and how much detail to include about the illness. Others were concerned that they were uncertain of their ability to respond and/or handle questions that would arise after disclosure. During the in-depth interviews, four of the 15 caregivers elaborated on this lack of confidence in their HIV knowledge.

> *“My problem was that I was not going to be able to face the questions that would arise and did not have a plan as to how I go about starting the conversation with him” (caregiver of participant 018, interview)*.

#### 1.2 Fear

Fear was a common reason for not disclosing in 40% of the caregivers (12/30). This was confirmed during the in-depth interviews. Fear of stigma, rejection from their child and fear in itself were equally reported.

Ten caregivers expressed fear of stigma. Five (50%, 5/10) had not disclosed prior to the study because they were concerned about how others would react to the child disclosing their status; another two (20%, 2/10) expressed concerns about how family would react to finding out that the child was HIV positive; and three (30%, 3/10) were afraid of the child being unable to maintain confidentiality (“keep the secret”) about their status.

Fear of social stigma was also evident during the in-depth interviews where one of the caregivers stated:

> *“We were scared and afraid that he would go about talking about it in the neighbourhood. Scared of being stigmatised” (caregiver of participant 028, interview)*.

Furthermore, five caregivers (17%, 5/30) cited a fear of rejection from their children after disclosure. During the in-depth interviews, this fear of rejection was not only limited to the caregiver being rejected by the child but also to the child being rejected by other family members

> *“I was scared to tell him he is HIV+ because it is only him that is infected … I thought that if the other two knew he was HIV they might distance themselves from him and not allow him to be part of them” (caregiver of participant 031, interview)*.

Generalised fear was observed in three caregivers (10%, 3/30), related to how the child would react to being disclosed to (i.e. running away or responding negatively to the news). Two other caregivers (7%, 2/30) were afraid to broach the subject of HIV and disclosure. This was reiterated during the in-depth interviews where the caregivers spoke of being afraid to broach the subject and one expressed concern that disclosing to the child would lead to worse consequences.

> “*How would I explain to him what is HIV? And what if he gets angry and kills himself” (caregiver of participant 005, Interview)*.

#### 1.3 Lack of Emotional Readiness

Guilt was the most common emotion preventing disclosure with caregivers describing feelings of guilt at having infected their children with HIV. This was evident during the in-depth interviews where some of the caregivers expressed their guilt and/or shame around their children’s status:

> “*Because I feel ashamed when I look at her always neh. And I was having that thought how am I going to tell her*” (*caregiver of participant 006, Interview*)

However, anxiety was only overtly observed in one caregiver (3%, 1/30) who appeared nervous about the child learning of her status.

Almost 10% (9/30) of caregivers also felt the need to be emotionally prepared before disclosing to their children. This ranged from one caregiver not feeling “ready” to broach disclosure to another who was still struggling with her own HIV diagnosis. Two other caregivers had only recently discovered their child’s HIV status when their child had been admitted to hospital for illness. These caregivers were not emotionally prepared to address disclosure with their children and later discontinued the study.

### 2. Reasons why caregivers chose to disclose through the intervention

During the in-depth interviews the caregivers were asked what prompted them to participate in the Disclosure study. Two sub-themes were evident when analysing their answers:

2.1 To explain why the child was taking medication
2.2 The need for assistance in disclosing and normalising the situation

#### 2.1 Explanations for why the child was taking medication

All participants in the disclosure study were on ARV treatment with the majority of the participants having been on treatment for most of their lives. Out of the 15 caregivers interviewed, nine (60%) expressed that their children were asking why they were taking medication.

> “*The reason I brought him to disclosure is that maybe he will get information about why he has to drink the pills and further tell him his HIV status*” (*caregiver of participant 005, interview*)

The lack of disclosure had led to caregivers misleading their children about the necessity for the medication. Children were told that they were taking medication for ailments such as Asthma, Bronchopneumonia, TB, Bronchitis, Tonsillitis and for the flu. Additional reasons included that they were health supplements and vitamins which would keep them from falling down often, getting injured, and to help them grow bigger and/or stronger. The caregivers also commented that at times their children would not adhere to the medicine regime and “lie” about having taken their medication

> “*she would give me stories/excuses when she was supposed to take her medication*” (*caregiver of participant 018, interview*).

#### 2.2 The need for assistance in disclosing and normalising the situation

Seven of the 15 (47%) caregivers interviewed expressed a need for assistance from more knowledgeable sources when disclosing to their children, as they, in particular the healthcare providers, would provide the necessary objectivity, support, information and have the ability to normalise the situation.

> “*I also needed some external help, if I can say so, to make him feel there is nothing wrong with whatever is happening or whatever is going on … I needed someone to assure him, besides me, to assure him that all’s good and that he’s good*.” (*caregiver of participant 001, interview*)
>
> By normalising the situation, the healthcare providers are able to put into words the fears and apprehensions the caregivers are experiencing. Through their understanding, support and ability to educate the caregivers, the caregivers are able to develop the ability to address disclosure with their children. Furthermore, the acceptance of status by both the healthcare providers and caregivers, as well as the knowledge that being HIV positive is not a death sentence or something to be ashamed about, assists the child in accepting their status.

### 3. The caregivers’ response to the Disclosure program (See Table 3)

Throughout the study, the healthcare providers recorded their observations of how the caregivers responded to the various stages of the disclosure program on the DCFs.

**Table 3:** Caregiver’s observed reaction to the Disclosure Process taken from the Disclosure Counselling Forms.

| Subtheme | Code | Examples from Data |
| --- | --- | --- |
| i. Pre- Disclosure | **Negative Responses** |  |
|  | a. Anxiety | “Father was told not to panic, and that he will be supported by the staff” (001, Week 24 Visit)<br>“Mom seemed nervous about the child knowing her status...Mom feels that the child will run away” (004, Week 12 Visit) |
|  | b. Hesitancy/ Avoidance | “The father said he is not ready to let the child know about HIV because he is not sure the child will comprehend the information” (001, Week 24 Visit)<br>“Mom has been avoiding any subject that related to HIV (002, Week 48 Visit) |
|  | **Positive Responses** |  |
|  | a. Happy | “mom says she's seen a big improvement in her child” (017, Week 12 Visit)<br>“Mom is happy with progress” (005, Week 12 Visit) |
|  | b. Satisfaction with program | “Father is happy about the process and promised to support the child” (001, Screening Visit) |
|  | c. Relief | “Mom relieved about the process” (014, Screening) |
| ii. Full Disclosure | **Negative Response** | “Granny was a bit anxious of her |
|  | a. Anxiety/ Uneasy | grandson's reaction and was also assured" (003, Week 48 Visit)<br>"mom was the one feeling uneasy specially after disclosing, she kept on doing disturbing mannerisms" (010, Week 48 Visit) |
|  | b. Counter-productive assurance | "Mom was in the session very anxious and at the same time trying to comfort her but a bit destructive" (015, Week 48 Visit) |
|  | **Positive Response**<br>a. Encouragement/Reassurance | "The mom really helped by encouraging her to talk" (011, Week 48 Visit)<br>"Tried to reassure him" (028, Week 48 Visit) |
| iii. Post -Disclosure | **Negative Response**<br>a. Emotionally distant | "Father seemed distant" (013, Week 72 Visit)<br>"No conversation after full disclosure as parents are not staying with the child...No one took the time to sit with her and talk, especially the mom (026, Week 72 Visit) |
|  | **Positive**<br>a. Satisfied/ Happy | "Mom very happy with her school progress and even at home she's coping very well" (006, Week 72 Visit) |
|  | b. Prompted further disclosure | "disclosure study helped her that she even decided to disclose to her older daughter cos (sic) she is HIV+ve too" (008, Week 72 Visit) |
|  | c. Relief | "mom was very anxious but she seemed <u>relieved</u> about her daughter after disclosure" (29, w48) |

#### Caregivers’ response Pre-Disclosure

The response was perceived by healthcare providers to be both negative and positive with six caregivers (6/30, 20%) reported to be anxious, and five (5/30, 17%) being hesitant. On the other hand, five (5/30, 17%) expressed their happiness and appreciation to be part of the process.

#### Caregivers response during Full Disclosure

Negative responses were more evident than positive ones with the most common during this period being anxiety or unease (6/30, 20%). One caregiver’s anxiety during this time appeared to be counter-productive during the session, having a negative impact, instead of being calming and encouraging, the caregiver’s own anxiety resulted in deflecting attention from the participant at a crucial stage of disclosure to that of the caregiver.

However, some caregivers responded positively, encouraging and reassuring the child.

#### Caregivers response Post-Disclosure

At their post-disclosure visits, the caregivers had more positive (23%, 7/30) than negative responses (7%, 2/30), including satisfaction with their child’s progress. Others reported relief that their children knew their status. One felt that the disclosure study was so helpful that she even decided to disclose to her other child. Other observations made included improved dyad relationships, understanding the importance of being responsible for the administration of medication and good nutrition. However, two caregivers (7%, 2/30) responded negatively, and were observed as being emotionally distant.

## Discussion

This qualitative investigation described the response of caregivers to the disclosure process aimed to assist them to disclose to their children’s HIV+ status to them. Caregivers indicated that, prior to the study, they were reluctant to disclose to their children due to feeling ill-equipped to address the subject, being afraid and not being emotionally ready. They were motivated to disclose during this study as medication adherence was becoming problematic and they felt more comfortable with a healthcare provider present to assist with disclosure and normalise the situation. Initially they presented as being both anxious and grateful for the study. As the study progressed to full disclosure, many caregivers reacted negatively with anxiety to the intervention. However, following full disclosure feelings of relief and satisfaction with their child’s progress was noted. This study also empowered a caregiver post-disclosure to independently disclose to her other child.

The caregiver’s attitudes towards HIV and HIV disclosure is shaped by their personal experiences within the environment and the attitudes of those around them, as surmised by social learning theory (27). This disclosure study allowed disclosure to occur in a contained environment under the nurturing and non-judgemental guidance of the healthcare provider. Healthcare providers modelled positive behaviours for both caregiver and child during disclosure, and guided the caregiver on how to disclose. This made the process less stigmatising, positive and open, and provided a safe space where trust was fostered. Perceptions that HIV should not be stigmatising and allowing caregivers space to gain confidence in their ability to manage disclosing to their children and also in talking about their own status were reinforced.

The healthcare providers’ ability to model healthy attitudes towards HIV and reduce negative emotions around disclosure, taught caregivers how to model the same behaviour for their children. This was especially evident in reflections from the caregivers where they dealt with their own HIV status during the disclosure study. By applying social learning theory, addressing the negative attitudes and stigmatization held within the wider community the study could shift the caregiver’s experience of their own disclosure, reduce their anxiety and shame, and thus encourage more positive responses towards HIV self-disclosure. Through this, children may learn not to fear the disease, and have less adverse responses when disclosed to (12, 39, 46).

Initially, many caregivers responded to the intervention with anxiety, concern that their children were too young for disclosure, fear of negative consequences, as well as a lack of confidence in their own knowledge of HIV. This is consistent with other studies (10, 23, 26, 41-45), indicating a need to address the stigma still highly present in our communities that contributes to secrecy around HIV. The fear of stigma from others also leads to self-stigmatisation, making it difficult for caregivers to accept their own diagnosis, and in turn projecting that unacceptance and stigma onto their child (13, 23, 39). This emphasises the need to equip healthcare providers to prepare and support caregivers throughout the disclosure process (27, 28, 43, 44, 47).

Previous research cited the caregivers’ fear of disclosure having a negative impact on children’s mental health but once disclosed to, the child was described as having lower anxiety and depression scores (26, 29, 40, 41-46). Early research reflected the fear of dying, and the portrayal of HIV-related deaths in the media was seen as another barrier to disclosure (41, 42). However, medical advancements and wider accessibility to ART may explain why this was not seen in our study. Importantly, two caregivers only discovered their child’s status when the child was admitted to hospital for illness prior to joining the PHRU Wellness Clinic shortly before the study began. Consequently, both discontinued the study. This emphasises the need to ensure that caregivers feel supported and have accepted their own diagnosis and that of their child before disclosure. It also stresses a need for community education to reduce the spread of misinformation (13, 23, 39).

Non-adherence was the most prominent motivation to disclose in this study. Prior to disclosure, caregivers would mislead their children about the need for medication and deflect diagnosis to maintain adherence. However, caregivers noted that these strategies were losing effect and realised the need to disclose. Improved adherence was noted in several cases in our study. This with subsequent improvement in the child’s wellbeing is in keeping with previous findings (13, 39, 47-50).

The need to normalise the child’s HIV status was another prominent motivating factor for disclosure. This concurs with studies that reported that children deserve the right to know their status (47, 49). Caregivers in this study stated that they required the assistance of the more knowledgeable healthcare providers in disclosing and normalising HIV to their children. Similarly, other studies have found that most caregivers either preferred healthcare providers to disclose to their children, or required support from knowledgeable healthcare providers during disclosure (4, 47, 50). This emphasises the significant role healthcare providers play in the disclosure process and is in accordance with other literature and recommendations made by WHO.

Given the barriers experienced by the caregivers prior to disclosure, an initial negative response early in the disclosure process is not surprising. However, this sample of caregivers had an almost equal number of negative and positive responses towards disclosure initially. Post-disclosure found caregivers responding positively with satisfaction and relief to the process. A sense of improved confidence, HIV knowledge and caregiver-child relationships were observed. Healthcare providers played a critical role in supporting the caregiver during disclosure, alleviating their anxieties and building their confidence. Furthermore, improved communication also assisted caregivers in becoming more comfortable talking about their own HIV status. Similarly, another HIV intervention study reported caregivers felt that the program made equipped them better to answer questions and to handle disclosure (51, 52).

Limitations in this study included: 1) positive responses may reflect a sampling bias, as these caregivers were willing and open to participate in both the main study and the sub-study. This could indicate a comfort and familiarity with the HIV clinic staff as most had been attending the clinic prior to the study. 2) Caregivers may have also provided socially desirable answers during the in-depth interviews due to familiarity with the interviewers from interactions during regular clinic visits. 3) A wide range of emotional responses were also not captured in the DCFs, a limitation when using observational reports. However, the in-depth interviews that followed the disclosure program, did allow the researchers to gain further insight into some of the DCF observations. 4) This study has a predominant female perspective as most of the experiences and views were of female caregivers, and the DCF’s were completed and analysed by a female team. However, in many interventions mothers are often the caregivers accompanying the child to the HIV interventions. Regardless of these limitations, this study has added to the very limited literature regarding caregiver’s reactions during disclosure especially in South Africa where the largest population of people living with HIV reside.

Understanding the reservations and motivations caregivers have towards disclosure can make an impact in shaping future HIV disclosure interventions. Many disclosure studies have focused on the children not their caregivers. This study suggests the need to design interventions with support structures in place that assist the caregiver through the disclosure process during and following full disclosure to the child. This study also highlighted the need for caregivers to be educated on the benefits of disclosure and risks of non-disclosure. Moreover, positive behaviours and attitudes modelled by healthcare providers play a significant role in assuring the caregiver during disclosure, and can start alleviating the effects of stigma from the community.

## Data Availability

All data produced in the present study are contained in the manuscript

## Acknowledgements

We would like to thank Nonhlanhla Mazaleni for the support she provided to study participants and families and Dr Stacy-Lee Sigamoney for writing the first draft of the protocol; Right to Care Foundation, Dr Leon Levin and Dr Julia Turner for the informative workshop on the “Right to Care Mini Flipster Disclosure Tool for Adolescents Ages 12 and up”; the PHRU clinic staff: Faith Madiehe, Mirriam Mantwa Kunene and Nkata Kekana, for their care and guidance provided to our patients and caregivers during this study.

